# The impact of policy timing on the spread of COVID-19

**DOI:** 10.1101/2021.03.16.21253764

**Authors:** Moshe Elitzur, Scott Kaplan, Željko Ivezić, David Zilberman

**Affiliations:** Department of Astronomy, Univ of California, Berkeley, CA 94720, USA; Dept of Agricultural & Resource Economics, Univ of California, Berkeley, CA 94720, USA; Dept of Astronomy, University of Washington, Seattle, WA 98195, USA

**Keywords:** COVID-19, pandemic, exponential growth, growth hindering

## Abstract

We model COVID-19 data for 89 nations and US states with a recently developed formalism that describes mathematically any pattern of growth with the minimum number of parameters. The results show that the disease has a typical duration of 18 days, with a significant increase in fatality when it lasts longer than about 4 months. Searching for correlations between “flattening of the curve” and preventive public policies, we find strong statistical evidence for the impact of the first implemented policy on decreasing the pandemic growth rate; a delay of one week in implementation nearly triples the size of the infected population, on average. Without any government action, the initial outburst still slows down after 36 days, possibly thanks to changes in public behavior in response to the pandemic toll. Stay-at-home (lockdown) was not the first policy of any sample member and we do not find statistically meaningful evidence for its added impact, similar to a recent study that employed an entirely different approach. However, lockdown was mostly imposed only shortly before the exponential rise was arrested. The possibility remains that lockdown might have shortened significantly the initial exponential rise had it been employed as first, rather than last resort.

## 1. Introduction

Various policies have been implemented to arrest the rise of the COVID-19 pandemic. To assess the impact of any given policy and compare its efficacy with other policies requires a description of the epidemic trajectory and objective measures of its trend. One approach is to construct detailed, dynamic epidemiological models and search for the impact of implemented policies on the model time variation (Brauner et al., 2020; Hsiang et al., 2020; Lai et al., 2020). An alternative is to employ a general mathematical description with the minimal number of free parameters and identify deviations from purely exponential growth. This is the approach we take here. We construct descriptive models of the pandemic first wave in a large sample, determine for each the point where the pandemic growth starts slowing down and look for statistical correlations with policy implementation dates.

## 2. General Description of Growth Slowdown

We have recently developed a formalism to describe any growth pattern with a time-varying growth rate (Elitzur et al. 2020; hereafter EKZ20). A brief summary of its main ingredients: Consider some quantity *Q* (> 0) such as national GDP, cumulative number of COVID-19 infections, etc., whose long-term variation with time *t* meets two criteria:

1. *Q* is monotonically increasing, so that its growth rate *g* = *d* ln *Q*/*dt* is ≥ 0.
2. *g* remains finite when *Q* → 0, that is, the limit *g*_u_ = *g*(*Q* → 0) is finite.≤

These conditions ensure an initial exponential growth phase where *Q* = *Q*_0_ exp(*g*_u_*t*), with *Q*_0_ an initial value. Subsequent growth may slow down for whatever reasons, an effect we termed *hindering*, so that *g*(*t*) ≤*g*_u_. Then the general solution of the equation of growth for *Q*(*t*) can be written as

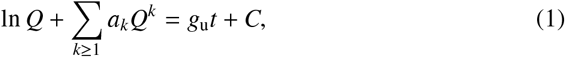

where *a*_*k*_ (≥0)^1^ are expansion coefficients that describe the hindering (slowdown) effect and *C* is an integration constant that ensures *Q*(*t* = 0) = *Q*_0_. Purely exponential growth occurs when *Q* is sufficiently small that the logarithmic term dominates the left-hand-side of eq. 1; this is the unhindered-growth regime with the unhindered growth rate *g*_u_. When the algebraic terms dominate, growth slows down and *Q* increases only as a power law—the hindered-growth phase. When the *k*-th order hindering term dominates, *Q ∼t*^1/*k*^; the higher is the hindering term the slower is the rise of *Q* with *t*.

Just as the Fourier series provides a generic description for all periodic phenomena, eq. 1 provides a generic description for any growing quantity that meets the two conditions listed above. This is the case even if *Q* displays occasional deviations from these requirements; for example, national GDP may suffer sporadic periods of negative growth, contracting during occasional recessions, but is still described by eq. 1 so long as its long-term behavior is one of growth. Every growth pattern can be described with a suitable set of hindering coefficients *a*_*k*_. A finite number of terms yields unbounded growth, with *Q* → ∞ at a continually decreasing growth rate, while infinite power series describe bounded growth, in which *Q* approaches asymptotically some upper limit.

With this formalism we successfully described the time variation of GDP and population in the US and UK, two nations with more than 200 years of continuous data coverage (EKZ20). In each of these cases the deviation of long-term growth from a pure exponential required no more than a single hindering term; there was no significant gain from adding more terms.

## 3. Hindering Description for the Covid-19 First Wave

Pandemic growth fits naturally into a hindering description. Typically, the number of infections grows exponentially at first, followed by a “flattening of the curve” as the growth slows down from its initial rate. These two stages are, respectively, the unhindered and hindered growth phases described above. The connection with standard epidemiological modeling is established in Appendix A.

We employ eq. 1 to model separately reported cumulative numbers of COVID-19 infections and deaths, denoted *Q* in each case. Most locations have already experienced a second and third pandemic wave in which the decline in the growth rate of *Q* is reversed. Describing such time variation would require a large number of expansion terms in eq. 1, some of which may need to be negative (see footnote 1). While this is possible in principle, our aim here is not to perform modeling for the sake of it but to gain insight into the impact of public policy on the growth of the pandemic. To enhance the analysis reliability, we simplify matters to the extent possible by restricting modeling to the pandemic first wave. The first waves of infections and deaths need not overlap. In fact, if every COVID-19 infection and death were detected and reported, the reported death counts would lag behind the cases by the duration of the disease. In practice, less-than-perfect detection and reporting afflict both datasets, especially the case counts. We identify the boundaries of each first wave from the behavior of the daily counts (*dQ*/*dt*). The first wave starts when the daily counts display unambiguously a monotonically increasing trend. With the non-parametric Mann-Kendall test (Appendix B.1) we determine the first occurrence of such a trend at the 95% confidence level in a consecutive sequence of daily counts. The first-wave endpoint is taken as the first minimum of the daily counts after the end of the initial exponential rise. To determine this minimum, we smooth the daily counts data with a simple 7-day moving average to avoid the impact of large fluctuations and the resulting outliers. Following this selection we fit the data for the pandemic first-wave with eq. 1 by varying the free parameters to minimize the residual sum of squares (RSS) of the cumulative counts *Q*. Technical details of the fitting process are described in Appendix B.

As an example of our analysis, Figure 1 shows the detailed results for New York State, one of the hardest hit locations in the early days of the pandemic. The data are shown as dots, with the case counts in the left column, the death counts on the right. Top panels show the cumulative counts, displaying a similar pattern: An initial exponential rise followed by “flattening of the curve”. The more moderate behavior during the latter phase is better discerned in the insets, which zoom in on the second half of each dataset with a linear, instead of logarithmic, *y*-axis. This expanded view also brings out problems in the data. The abrupt dislocation on day 106 (June 30) of the death counts arises from changes in reporting protocols. The bottom panels show the daily counts, whose overall trends are conveyed by the displayed 7-day moving averages. The extreme outlier on day 106 of the daily death counts causes a seven-day upward displacement of the moving average and the dislocation in the corresponding cumulative count.

**Figure 1:**
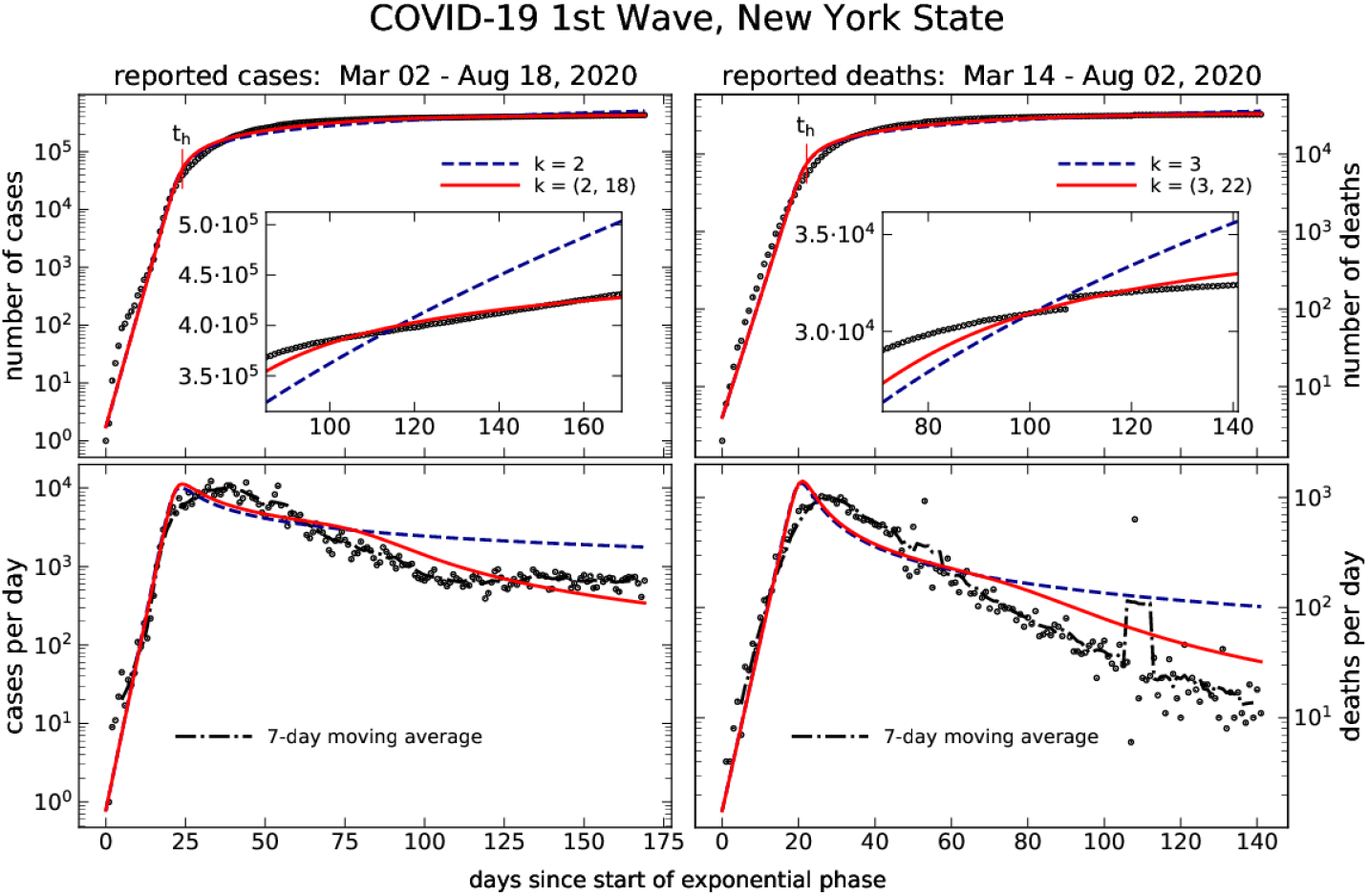
The Covid 19 pandemic first wave in New York State. Left column shows reported cases, right column reported deaths. Dots show the data, lines are best fits with eq. 1 for single- and two-power models, as labeled. *Top panels*: Cumulative number from start of the exponential phase; this phase ends in the transition to hindering, marked *t*_h_ (see text for details). The insets zoom-in on the second half of each dataset with linear *y*-axis instead of logarithmic. *Bottom panels*: Daily counts; dot-dashed line is the 7-day moving average. The fitting was done for the cumulative counts (*Q*; top panels). The bottom-panel curves show *dQ*/*dt* and involve no fitting, fully determined from the models in the top panels.

Lines show our models, obtained by fitting eq. 1 to the cumulative counts (top panels). The point where the model-calculated growth rate *g* decreases to half its initial value, the unhindered *g*_u_, is marked *t*_h_. This is where the deviation of *g* from *g*_u_ becomes significant, an indicator of the end of the initial exponential phase and the transition to hindered growth. For the most part, the data points are barely distinguishable from the model plots in both top panels. The quality of the fits is further illustrated by the fraction of variance unexplained (FVU = 1 − *R*^2^ where *R*^2^ is the coefficient of determination), which is only ∼ 1% in both cases. Although the curves for single-term models are hardly distinguishable from their 2-term counterparts, the additional hindering term of the latter does improve the fit for *Q*. The *F*-test null hypothesis that the coefficient of the 2nd term vanishes (see Appendix B.2) can be rejected at high confidence levels: more than 99% (p-value of 2.5·10^−7^) for the case counts and 98% for the death counts. The impact of the second power-law becomes evident in the insets, and stands out prominently in the lower panels, which show the daily counts. While the best-fitting single-power models describe properly the growth initial slowdown, they are clearly inadequate for the subsequent steeper falloff.

Adding more hindering terms would further improve the fits, at the risk of over-fitting and chasing noisy structure in the data. This we wish to avoid as we are only interested in capturing meaningful long-term trends. Two hindering terms successfully achieve that for the displayed datasets. Importantly, although the single-term models do not properly describe the late hindering stages of the first wave, they are practically indistinguishable from the two-term solutions during the exponential rise and early hindering phase. The unhindered growth rates *g*_u_ are the same to within a fraction of a percent, the hindering times *t*_h_ differ by less than a day.

The plots of *dQ*/*dt* involve no fitting. They are fully derived from the models for *Q*(*t*), shown in the upper panels. Since daily counts are frequently noisy and irregular, modeling the cumulative counts is a significant advantage of our method. The cumulative counts are produced in a running sum, effecting a simple smoothing that preserves the underlying trends as evidenced by the fact that the long-term variation of *dQ*/*dt* is captured reasonably well by our models for *Q*.

## 4. Policy Impact

A variety of factors can play a role in arresting the exponential rise of the pandemic, many of them unknown. The one we wish to study is the potential impact of policy decisions, such as business closures, lockdown (stay at home), etc. The approach we take here is: (1) Construct hindered-growth models of the first wave of the pandemic in a sample of locations, (2) determine from them the observable *t*_h_, the onset of hindering, for every sample member, and (3) look for correlations with policy implementation dates, the independent variables.

We study five different policy categories, which are the most commonly used around the world in attempts to slow the growth of the pandemic: Educational facility closures, essential and non-essential business closures, travel restrictions, restrictions on gatherings, and stay-at-home orders. With data on policy implementation from the Institute of Health Metrics and Evaluation (IHME)^2^ and Brauner et al. (2020) we selected 52 countries and 47 U.S. states. All 99 entries on this list instituted at least one of the aforementioned policies.

### 4.1. Modeling and results

The key quantity in assessing the potential impact of a policy in slowing the growth of the pandemic is the length of the exponential phase—same as the onset day of hindering *t*_h_. We determine this parameter by modeling the COVID-19 data of our sample members^3^ with a single hindering term in eq. 1. As shown above, single-power hindering yields reliable estimates of *t*_h_. With a single hindering term, our models modify pure exponential growth with the minimal number of parameters: the power *k* and coefficient *a*_*k*_.

Excessive irregularities in the daily counts of some locations caused difficulties to our data selection algorithm, which hampered modeling of those data sets. We ended up with 89 reliable models for case counts and 81 for death counts. Detailed tabulations of our data and model results are provided in Appendix C. Figure 2 presents histograms of key results. Panel (a) shows that the first wave lasts, on average, a full month longer for deaths than for cases. Both histograms peak at around 90 days, but the deaths histogram peaks again around 160 days and has an extended tail. In contrast, panel (b) shows that the length of the exponential phase, which starts with the onset of the first wave and ends *t*_h_ days later, is essentially the same for both cases and deaths. This can also be deduced from panel (c): deaths lag behind cases by the same amount, on average, for both the starting date of the first wave and the hindering date; the exponential phase of COVID-19 deaths is simply a shift of the cases by about 18 days. The implication is that the much longer duration of the deaths first wave arises from the hindered phase, indicating that beyond a certain threshold (about 120 days), the death probability increases significantly with the length of the illness. Finally, panel (d) shows that case counts rise faster than death counts during the unhindered phase; the unhindered growth rate for case counts is ⟨*g*_u_⟩ = 34.9% per day, where angle brackets denote the distribution mean, for death counts it is 23.3% per day. As a result, the deaths:cases ratio, also known as the observed case-fatality ratio, is decreasing with time. Such a decline may be triggered by increased testing, which makes the number of reported cases rise faster than actual infections, as well as improvements in medical treatment of the disease, which decrease the fraction of deaths.

**Figure 2:**
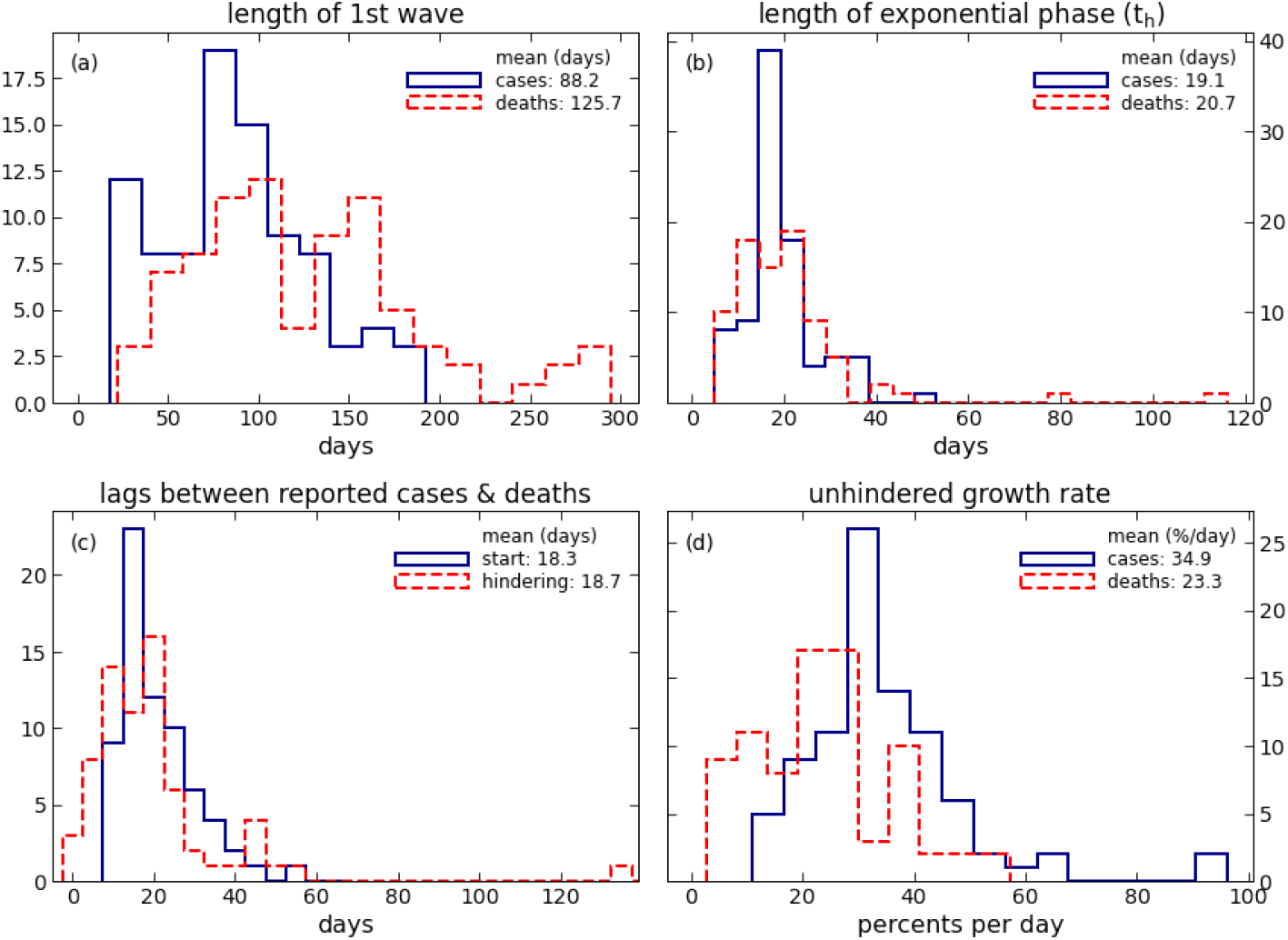
Histograms of the length of the COVID-19 *(a)* first wave and *(b)* initial exponential (unhindered) phase (same as *t*_h_, the onset day of hindering). *(c)*: Lag times between reported cases and deaths for the start of the 1st wave and the onset of hindering, as labeled. *(d)*: Histograms of the initial, unhindered growth rate *g*_u_.

Extraneous factors decrease the observed case-fatality ratio during the first ∼70 days, yet that ratio rises significantly after ∼120 days. This enhances confidence in the robustness of the conclusion that the late rise reflects an increase in the intrinsic fatality of the disease when it lasts longer than ∼4 months or so.

### 4.2. Correlations

Public policy is effective in controlling the pandemic if it shortens *t*_h_, the length of the exponential phase. The policy history of each location is characterized by the ordered sequence of implementation dates for each of the five policy categories described above, resulting in 120 possible combinations. Since this exceeds the number of our data points, we collapse the space of independent variables by considering at every location only the first policy implemented, whatever it is, and test for the correlation of its implementation day *t*_policy1_ (days after start of the exponential phase) with *t*_h_. This is done with simple regression analysis for the case counts; there is no need for a repeat with the death counts because, as shown above, the exponential phase of COVID-19 deaths is, on average, a simple time shift of its case counterpart.

Regression analysis for the observable *t*_h_ and independent variable *t*_policy1_ yields a highly significant linear correlation (p-value = 1.09·10^−4^). The results are tabulated in the first column of Table 1 and shown in panel (a) of Figure 3 together with the data points. On average, the first policy was implemented 5.91 days into the exponential phase, hindering occurred 13.21 days later. The point on the correlation line with *t*_policy1_ = 29.1 days has *t*_h_ = *t*_policy1_, which would imply that the first policy was imposed on the same day hindering was setting in. This point is beyond the range of our sample—the latest implementation of a first policy, by the United Kingdom, was on day 24 of the exponential phase.

**Table 1:** Correlations between length of exponential phase and policy implementation

| sample<br>size | $t_h = a_1 t_{\text{policy1}} + \text{const}$ | | | $+ a_2 \Delta_{\text{lock}}$ |
| --- | --- | --- | --- | --- |
|  | full<br>89 | no lockdown<br>29 | lockdown<br>60 | lockdown<br>60 |
| $a_1$ | 0.43 (0.11) | 0.35<sup>†</sup> (0.20) | 0.47 (0.12) | 0.51 (0.12) |
| $a_2$ | — | — | — | 0.21<sup>‡</sup> (0.16) |
| const | 16.59 (1.00) | 15.49 (2.05) | 17.02 (1.07) | 14.59 (2.15) |

*Notes:* Regression coefficients for $t_h$ , length of the exponential phase, on $t_{\text{policy1}}$ , implementation day of first policy, and $\Delta_{\text{lock}}$ , the additional number of days to lockdown. Numbers in parentheses are the standard errors. All p-values are $\ll 0.01$ unless noted otherwise.
<sup>†</sup>The p-value for this entry is 0.10
<sup>‡</sup>The p-value for this entry is 0.20

**Figure 3:**
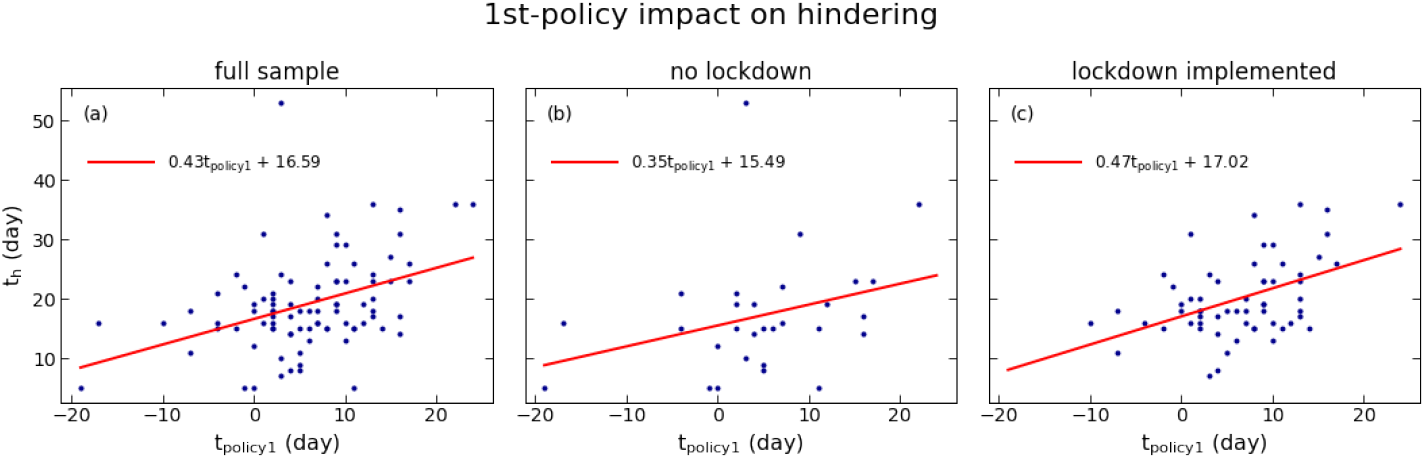
Variation of the onset of hindering, *t*_h_, with the first implementation of a policy, *t*_policy1_, during the first wave of COVID-19 case counts. Results for *(a)* the full sample, *(b)* the subsample of the entities that did not implement lockdown and *(c)* those that did. The zero-point of every axis is the start of the initial exponential phase. Dots show the data, straight lines the results of linear regression analysis (see Table 1).

The zero-intercept of the correlation line, 16.6 days, is the mean length of the exponential phase when the first policy is implemented right at the start of this phase. Twelve locations took action even earlier, which does not seem to have brought additional benefits: the mean value of *t*_h_ for this group is 15.3 days, no different than the zero-intercept within the standard error of 5.97 days. Apparently, the benefits accrued by early policy implementation are not improved further by acting before the start of the exponential phase.

The correlation shows that a delay of one week in implementing the first policy adds, on average, 3 days to the continuation of exponential rise at its initial growth rate (34.9% per day), increasing the cumulative case number by a factor of 2.85.

### 4.3. Lockdown’s potential impact

Stay at home (lockdown) is the most restrictive of the policies implemented by members of our sample. It was imposed by 60 of the 89 sample members, but never as the first policy; in fact, it was the last one in all but 12 cases. As a result, the *t*_h_–*t*_policy1_ correlation just derived does not include lockdown direct effect, only its potential added impact on top of other policies that were implemented earlier. To assess the potential added impact of lockdown, we split the sample into those that did and did not implement lockdown; the data points for each subsample are shown in panels (b) and (c) of figure 3. First we tested whether the two subsamples come from different distributions—if lockdown had no impact on *t*_h_, their *t*_h_ data would have been drawn from the same parent distribution. This hypothesis is rejected by the Kolmogorov-Smirnov two-sample test with a p-value of 0.05, and by the Mann-Whitney rank test with a p-value of 0.01. The two subsamples come from intrinsically different distributions, but that does not establish a direct correlation between *t*_h_ and lockdown. For that we repeated the *t*_h_–*t*_policy1_ regression analysis for each subsample separately. The resulting correlations are listed in Table 1 and plotted in Figure 3. The correlation is statistically significant for the locations that did implement lockdown (*p* = 1.84·10^−4^), but not for those that didn’t; their *t*_h_–*t*_policy1_ distribution is statistically indistinguishable from random scatter, differing from it by less than 2 standard deviations. At the same time, the *a*_1_ coefficients estimated from the two subsamples are also statistically indistinguishable from each other. There is no definitive outcome to the comparison of these two subsamples.

Attempting to tease out the potential added impact of lockdown, we are forced to rely solely on the subsample of 60 locations that did impose it. Denote by Δ_lock_ the number of days that passed from the implementation of the first policy (*t*_policy1_) to lockdown; Δ_lock_ ranges between 1 and 22 days, with a mean⟨Δ_lock_⟩= 10.58 day. There is a small but significant (*p* = 0.05) negative correlation between the variables with slope −0.20; that is, a 5-days delay in imposing a first policy hastened the subsequent implementation of lockdown by a day. Adding Δ_lock_ to the regression analysis as a second independent variable yields the results listed in the last column of Table 1. The coefficient *a*_2_ is 0.21 with a p-value of 0.20, thus there is no statistically significant evidence for the impact of Δ_lock_. However, lack of decisive statistical evidence does not prove that lockdown had no impact. The 95% confidence interval for *a*_2_ is [-0.11, 0.52], suggesting that approximately 90% of the distribution of its estimate is > 0.^4^ It must be noted, too, that lockdown was implemented, on average, only 3.03 days before the onset of hindering. By that time, the first policy had already run 78% of its course. Moreover, 19 of the 60 sample members, almost a third, imposed lockdown only after *t*_h_, too late to make any impact the onset of hindering. The possibility that lockdown might have shortened significantly the exponential phase had it been employed as first resort instead of last remains open.

## 5. Summary and Discussion

In the first part of this study we construct phenomenological descriptions for the COVID-19 data of numerous nations and US states, in the second we conduct statistical analysis of the derived model parameters in search of evidence for the impact of government policies on the pandemic growth trajectory. The phenomenological modeling is aimed at getting an objective, reliable measure of “flattening of the curve”, a day that marks consistent decline of the growth rate from the initial value it had during the exponential outburst of the pandemic. Our recently developed hindering formalism (EKZ20) identifies the desired parameter as the hindering time *t*_h_. We determine *t*_h_ from fits to the data done with minimal modification to pure exponential growth, adding a single power-law term to the modeling function (eq. 1). The simplicity of this method helps reduce the risk of potential pitfalls and makes it easy to study a large sample that contains every nation and US state with relevant data. Modeling cumulative, rather than daily, counts is another advantage of our method since daily count data are frequently rather noisy, containing many spurious zeros; a standout example is Sweden, which does not even report COVID-19 cases and deaths every day of the week. While fitting cumulative counts, our models also capture the variation of daily counts reasonably well, as evidenced by the example of NY State (figure 1).

With a sizeable sample, our models uncover some interesting properties of the COVID-19 disease itself, showing that the mortality rate varies with the sickness duration (figure 2). Other than an 18–19 day lag, the main segments of the histograms for death and case counts are similar, indicating that the mean duration of the disease is ∼ 18 days; after that, infected people either recover or die. This pattern lasts for ∼ 70 days. The mortality rate rises for those that remain sick longer, with a significant increase when the illness lasts for more than ∼ 4 months.

The end of the pandemic initial exponential rise is controlled by numerous factors, many of them unknown or unmeasurable. Here we searched for the potential impact of the governmental policies implemented by our sample members. Linear regression analysis yields a highly significant correlation between the implementation day of the first policy, whatever that policy is, and flattening-of-the-curve day *t*_h_. Delaying the former by one week almost triples the cumulative case number, on average. However, policy makers do not have all of the relevant information when making decisions, and governmental policy is not the only factor controlling the pandemic trajectory. In twelve locations, the first policy was implemented before the start of exponential rise, yet their average *t*_h_ is within a standard error of the zero intercept of the correlation we find; that is, implementing the first policy on or before the first day of the exponential rise limited its duration, on average, to 16–17 days, but not less. This minimal length may reflect a combination of factors, including the virus incubation period, societal response, etc. At the other end, our correlation implies that the initial exponential phase would end just as the first policy is imposed if the latter were delayed until day 29; namely, the initial growth would start slowing down after a month even without governmental action. While no government waited that long, Denmark and South Korea enacted their first policy after the initial exponential rise had already come to an end and 13 other entities acted within 5 days of that end point. Yet except for one extreme outlier (Nebraska, with *t*_h_ = 53 days), the length of the exponential phase never exceeded 36 days, within the errors of our correlation’s prediction. A possible explanation for why uncontrolled growth slows down on its own after 36 days is a change in public behavior without government action, triggered by the severity of the pandemic toll.

Stay at home (lockdown) is excluded from the evidence we find for the impact of the first implemented policy. Because of reluctance to employ it, this severe restriction was generally imposed as a last resort (never the first), too late to do much good; policy makers did not realize that the initial exponential rise was about to end, or even already ended, thanks to the less restrictive measures implemented earlier. There is a small negative correlation between Δ_lock_ and *t*_policy1_, namely, the longer decision makers waited to implement their first policy, the less time they took subsequently to impose lockdown. We conducted separate analysis in search of an added impact of lockdown and did not find statistically significant evidence for it.

Two recent studies serve as important benchmarks for our results. The one by Brauner et al. (2020) constructed detailed epidemiological models of 41 nations, all included in our sample, and reached the same conclusions—strong evidence for the impact of the first policy but not of lockdown. It is encouraging that we independently find the same results with such widely different approaches. However, despite the lack of decisive statistical evidence for an added impact from the late implementation of lockdown, a close examination of our regression results shows that the possibility of such impact cannot be rejected. It remains entirely possible that lockdown might have shortened significantly the initial exponential phase had it been employed as first, rather than last resort. The study by Hsiang et al. (2020) extensively examined the impact of policy implementation on the evolution of the COVID-19 pandemic at the local, regional, and national levels in six large countries. Their findings suggest that in the absence of policy action, early growth rates in these nations averaged approximately 38%; by comparison, we find an average unhindered growth rate for case counts of 34.9%, well within a single standard error of their estimate. Along with estimating the aggregate impact of all implemented policies on the COVID-19 growth rate by country, they estimate individual impacts as well. These support our findings that “home isolation” (i.e. lockdown) had a lesser impact on slowing the growth than other notable policies implemented earlier on, including emergency declarations, school closures, and other social distancing measures. In the six nations they study, home isolation was never the very first policy implemented.^5^ It is clear that when extending the sample of analyzed entities to the 89 we examine here, the timing of policy implementation was a more important factor in inducing hindering than the strictness of the policy.

Epidemiology models solve detailed rate equations specific to an epidemic to describe its growth trajectory. Such approach is essential for modeling of individual countries and for gaining insight into the various factors affecting the pandemic spread. The phenomenological description presented here is not specific to COVID-19, in fact to any epidemic—the same hindering function (eq. 1) was also used to fit time variation of GDP and population (EKZ20). Hindering is a generic description of any growth pattern just as Fourier analysis is a generic description of all periodic phenomena. Although such phenomenological modeling does not provide insight into the mechanisms driving the growth, it is a useful method to concentrate on the minimal number of parameters most relevant for the problem at hand, model large samples and classify growth patterns, providing a practical, complementary tool to guiding policy decisions.

## Data Availability

Research based on public data. All data sources are referenced in the article.

https://github.com/CSSEGISandData/COVID-19

https://github.com/nytimes/covid-19-data

## Acknowledgments

SK and DZ acknowledge support from IGI and NSF CBET (Grant no. 2030362)

## Appendix A. Epidemiology Models and Hindering

Our hindering formalism can describe any growing quantity *Q*, irrespective of the underlying processes driving its growth. The connection with such processes can be done through the growth rate *g* = *d* ln *Q*/*dt* and its relation to the parameters of a dynamic model for the growth of *Q*. A key concept in epidemiology modeling is the basic reproduction number of an infection, *R*_0_, defined as the expected number of secondary infections generated by an average infectious case in a fully susceptible (uninfected) population. This quantity determines the potential for an infectious agent to start an outbreak, the extent of transmission in the absence of control measures, and the ability of control measures to reduce spread. The effective reproduction number *R*_t_ (also denoted *R*_*e*_), is the number of infections directly generated by a single infectious individual at time *t* after the outbreak, and thus is applicable to an ongoing epidemic. Since *R*_t_ is dimensionless while *g* is rate, relating the two requires an independent time scale that characterizes the epidemic.

The spread of infectious diseases is traditionally described with compartmental models—the population is assigned to labeled compartments and rate equations describe the movement between them. In the most basic SIR models, S denotes susceptible individuals that have never been infected and I infectious ones (Miller, 2017). A susceptible individual that contracts the disease transitions to the I compartment and later removed into the R compartment as a result of recovery, and presumably developing resistance, or death. The resulting rate equation for the number of infectious individuals, those in the I-compartment, can be written as a growth equation, yielding

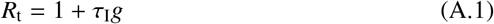

where τ_I_ is the mean infectious period (τ_I_ = γ^−1^ where γ is the transition rate from compartment I to R).

An infection can have a significant incubation period during which an individual has been infected but is not yet infectious himself. During this period the individual is in compartment E (for exposed), resulting in SEIR models (for Susceptible–Exposed– Infectious–Removed) which have been employed in numerous studies of COVID-19 (e.g. Annas et al., 2020; Brauner et al., 2020; He et al., 2020; Hsiang et al., 2020; Lai et al., 2020; Ma, 2020; Mwalili et al., 2020). If τ_E_ denotes the mean latent period, i.e., time from infection to onset of infectiousness, the sum τ_I_ + τ_E_ is called the *serial interval* (Lipsitch et al., 2003). A common growth rate for the *E*- and *I*-fractions can be obtained from a solution of an eigenvalue problem. The result is the quadratic relation

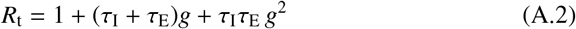

(Lipsitch et al., 2003). The limit τ_E_ = 0 (no latent period) reverts to the SIR model result (eq. A.1).

In these results, the growth rate *g* is that of *I*, the infection daily counts. The quantity that we model is *Q*, the cumulative case count. The hindering formalism (EKZ20) shows that the general description of *Q* can be written as (cf equation 1)

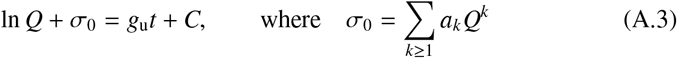

The free parameters *g*_u_ (the unhindered growth rate of Q), {*a*_*k*_} (the expansion coefficient set) and *C* (an integration constant that ensures an initial condition) are determined by best-fitting the data for *Q* with this expansion. The required growth rate *g* of *dQ*/*dt*, the daily counts, can then be derived through straightforward differentiation and algebraic manipulations:

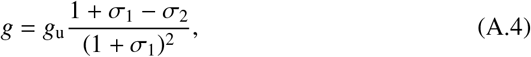

where

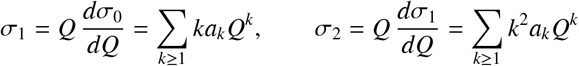

The effective reproduction number, and its time variation, can be obtained by inserting this result in eq. A.2.

## Appendix B. Data Analysis

Datasets of points *q*_0_, *q*_1_… at times *t*_0_, *t*_1_… are modeled with the function *Q*(*t*), defined in parametric form in eq. 1. Best-fitting models are obtained by minimizing the residual sum of squares (RSS) of the data and model points *Q*_*i*_ = *Q*(*t*_*i*_). Because of the large dynamic range spanned by typical datasets, we give all data points equal relative weights (σ_*i*_ ∝ *q*_*i*_) so that the minimization is performed on RSS = *i* (*Q*_*i*_/*q*_*i*_ − 1)^2^. It is important to note that we only seek the minimum of RSS; its actual magnitude is irrelevant (no need to specify the magnitude of the proportionality constant in σ_*i*_ ∝ *q*_*i*_).

### Appendix B.1. Mann-Kendall Test

The logarithmic term in eq. 1 describes pure exponential growth, corresponding to a constant growth rate. The first step is to determine whether any hindering corrections (the algebraic terms) need to be included in the model. Such terms describe slowdown of growth, thus the first step is to determine whether the data provide evidence for such slowdown, i.e., a decline in the growth rate *g*. To that end we compute *g* from the data with a finite-differences calculation and perform the Mann-Kendall (hereafter MK) test on its time series. This non-parametric test determines whether or not there is a monotonic trend in a given dataset. Its *Z*-statistic is computed from the signs of differences between data pairs.^6^ There are no assumptions regarding the distribution of data points and no requirement that the errors be normally distributed. The null hypothesis (H_0_) is no trend in the time series, in which case the test statistic *Z* is distributed according to the normal distribution with zero mean and unity standard deviation. Positive (negative) *Z* indicates an increasing (decreasing) trend. To determine the presence of hindering we test the null hypothesis against the alternative hypothesis (H_a_) that there is a downward monotonic trend (*Z* < 0) in a one-tailed test. The presence of hindering is established at the 99% confidence level when *Z* < −2.33, which can be achieved with as few as 6 data points. By example, the NY dataset analyzed in §3 yields *Z* = −16.1 for the case counts and − 18 for the death count, therefore in each case the null hypothesis can be rejected in favor of the alternative H_a_ with a p-value that is essentially zero within the numerics capacity. The MK-test provides robust statistical confirmation that neither dataset can be meaningfully described with pure exponential growth. Hindering corrections must be added in both cases.

### Appendix B.2. Hindering Terms; F-Test

When the modeling function must contain at least one hindering (i.e., algebraic) term, the question is what power-law to use for that term and how many additional ones might be needed. Our approach in EKZ20 was to handle this as a Taylor series expansion: start with *k* = 1 and add consecutive powers until reaching negligible marginal contribution. Here we adopt a different approach because of a significant difference between the two situations. As is evident from eq. 1, the independent variable is the dimensionless *x* = *g*_u_*t* = *t*/*T*_u_, where *T*_u_ = 1/*g*_u_ is the growth time during the unhindered phase. For both the GDP and population datasets of the US and UK, analyzed in EKZ20, the magnitude of *g*_u_ is less than ∼ 4% per year so that *T*_u_ is at least 25 years. With a time span of ∼ 200 years, *x* was at most ∼ 8 in those datasets. In contrast, *g*_u_ for the COVID-19 data is ∼ 30% per day so that *T*_u_ is ∼ 3 days and *x* typically extends to more than∼ 40. Because of the much larger range of *x*, proper modeling of these datasets could require a large number of terms to describe structures that may reflect noise rather than fundamental trends.

To avoid such potential pitfalls we devised an alternative approach that seeks to identify the long-term trends in the data rather than construct the absolute best fit. In the first step we model the data with a single hindering term and determine the power that provides the best fit; this ended up being *k* = 2 for the NY case count and 3 for the deaths (fig. 1). Next we employ a two-terms model whose first power is *k* of the best-fitting single-term model. The power of the second term is varied along *k* + 1, *k* + 2… until achieving the best fit with this two-term model. For the NY case counts the 2nd term ended having a power of 18, for death counts 22 (figure 1). Since the addition of a term can be expected in itself to improve fitting, we must determine the statistical significance of this improvement. The single-term model is a restricted form of the two-term model, with the coefficient of the 2nd term restricted to zero, thus the problem can be handled with the *F*-test, assuming that the unobserved error is normally distributed (Wooldridge, 2009).^7^ The null hypothesis is that the additional term has no effect on the dependent variable so that its coefficient should be zero. The number of data points, the ratio of RSS for the two models and their number of free parameters are combined to form the *F*-statistic (or *F* ratio); it follows an *F*-distribution, which arises as the ratio of two normal random variates. The *F*-statistic is compared with a critical value *F*_crit_, determined by the degrees of freedom for each model and an error level α. When *F* > *F*_crit_, the null hypothesis can be rejected at the confidence level 1 − α, the probability of a false rejection is less than α.

### Appendix B.3. Modeling of the Full Sample

The full sample was fitted with single-term models. This simplest form of hindering suffices to reproduce reliably the quantities of interest here—the hindering time *t*_h_, an objective measure of the “flattening of the curve” day, and the unhindered growth rate *g*_u_. Each dataset was also fitted with the logistic function (Griliches, 1957), which became the best overall fit when it produced a smaller RSS than the best-fitting hindering model. This was the case in a number of datasets for death counts. Since the regression analysis to determine the impact of public policy on the growth of the pandemic employed only the case counts, it did not involve any logistic models. The full results are tabulated below.

## Appendix C. Tables

Tables of our model results for case and death counts during the COVID-19 pandemic first wave are presented here in printed form and supplied separately in csv file format. Column headers are as follows:

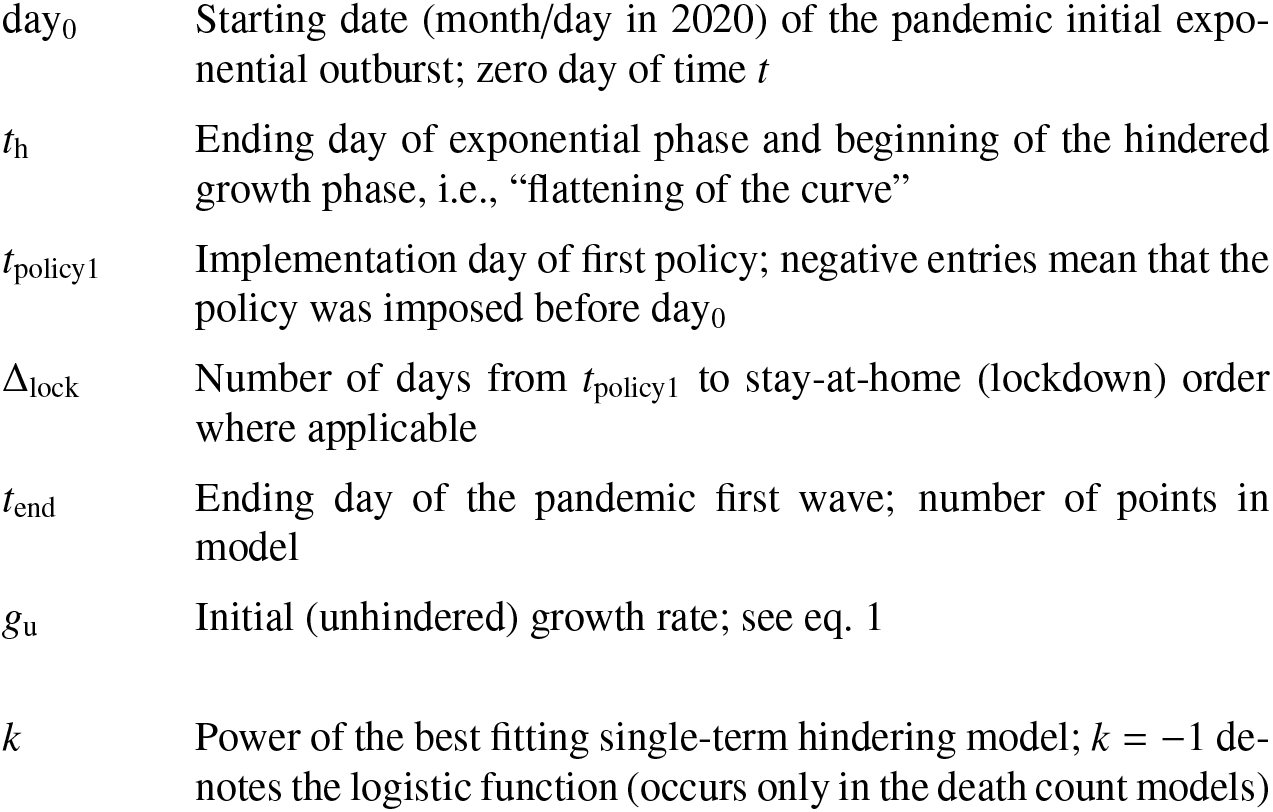

|  |  |
| --- | --- |
| $day_0$ | Starting date (month/day in 2020) of the pandemic initial exponential outburst; zero day of time $t$ |
| $t_h$ | Ending day of exponential phase and beginning of the hindered growth phase, i.e., “flattening of the curve” |
| $t_{policy1}$ | Implementation day of first policy; negative entries mean that the policy was imposed before $day_0$ |
| $\Delta_{lock}$ | Number of days from $t_{policy1}$ to stay-at-home (lockdown) order where applicable |
| $t_{end}$ | Ending day of the pandemic first wave; number of points in model |
| $g_u$ | Initial (unhindered) growth rate; see eq. 1 |
| $k$ | Power of the best fitting single-term hindering model; $k = -1$ denotes the logistic function (occurs only in the death count models) |

The columns for the policy data *t*_policy1_ and Δ_lock_ are entered only in the tables for case counts; they are not repeated in the tables for death counts.

**Table Appendix C.1:**
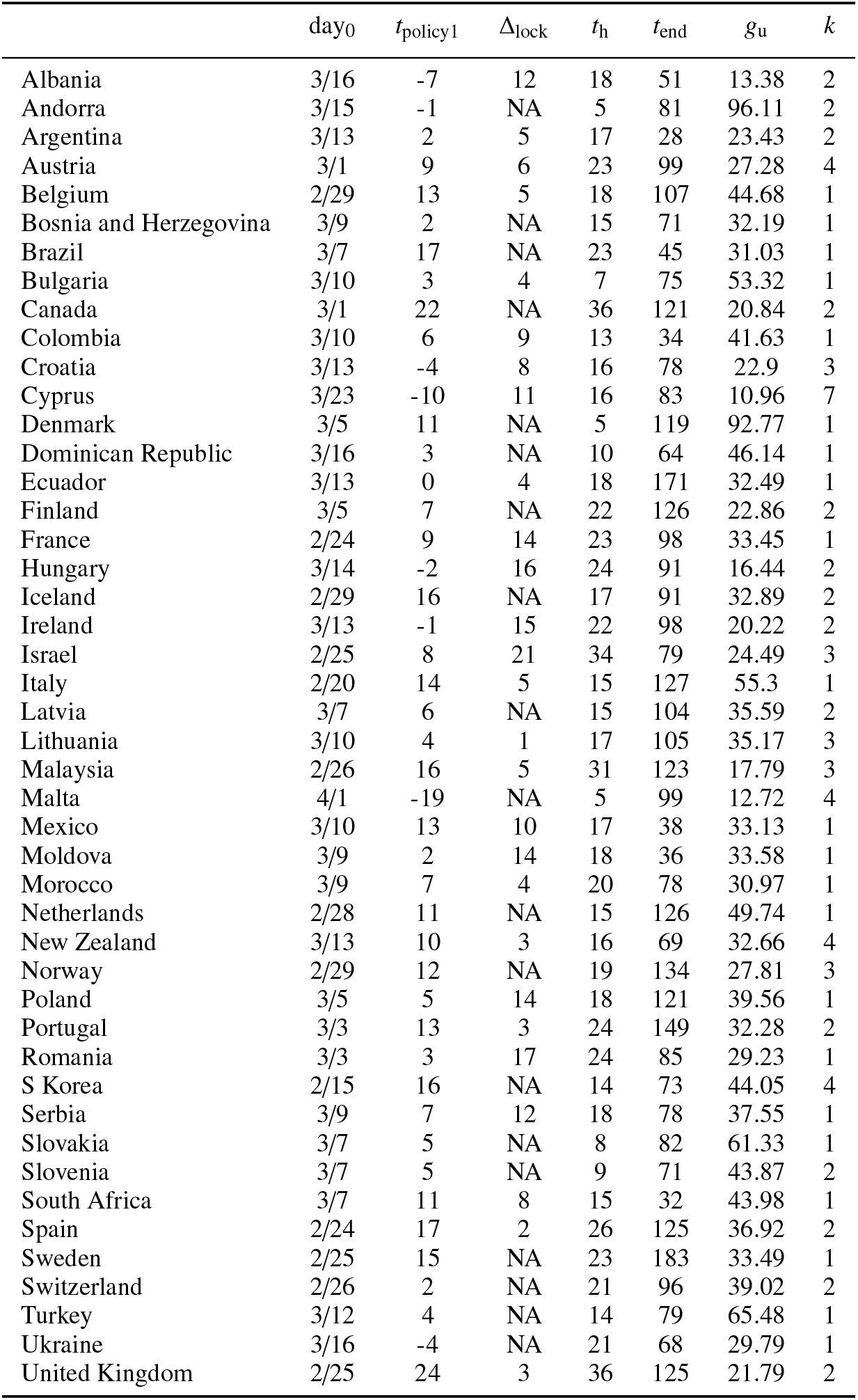
COVID-19 case counts; Nations

| | day<sub>0</sub> | $t_{\text{policy1}}$ | $\Delta_{\text{lock}}$ | $t_h$ | $t_{\text{end}}$ | $g_u$ | $k$ |
| --- | --- | --- | --- | --- | --- | --- | --- |
| Albania | 3/16 | -7 | 12 | 18 | 51 | 13.38 | 2 |
| Andorra | 3/15 | -1 | NA | 5 | 81 | 96.11 | 2 |
| Argentina | 3/13 | 2 | 5 | 17 | 28 | 23.43 | 2 |
| Austria | 3/1 | 9 | 6 | 23 | 99 | 27.28 | 4 |
| Belgium | 2/29 | 13 | 5 | 18 | 107 | 44.68 | 1 |
| Bosnia and Herzegovina | 3/9 | 2 | NA | 15 | 71 | 32.19 | 1 |
| Brazil | 3/7 | 17 | NA | 23 | 45 | 31.03 | 1 |
| Bulgaria | 3/10 | 3 | 4 | 7 | 75 | 53.32 | 1 |
| Canada | 3/1 | 22 | NA | 36 | 121 | 20.84 | 2 |
| Colombia | 3/10 | 6 | 9 | 13 | 34 | 41.63 | 1 |
| Croatia | 3/13 | -4 | 8 | 16 | 78 | 22.9 | 3 |
| Cyprus | 3/23 | -10 | 11 | 16 | 83 | 10.96 | 7 |
| Denmark | 3/5 | 11 | NA | 5 | 119 | 92.77 | 1 |
| Dominican Republic | 3/16 | 3 | NA | 10 | 64 | 46.14 | 1 |
| Ecuador | 3/13 | 0 | 4 | 18 | 171 | 32.49 | 1 |
| Finland | 3/5 | 7 | NA | 22 | 126 | 22.86 | 2 |
| France | 2/24 | 9 | 14 | 23 | 98 | 33.45 | 1 |
| Hungary | 3/14 | -2 | 16 | 24 | 91 | 16.44 | 2 |
| Iceland | 2/29 | 16 | NA | 17 | 91 | 32.89 | 2 |
| Ireland | 3/13 | -1 | 15 | 22 | 98 | 20.22 | 2 |
| Israel | 2/25 | 8 | 21 | 34 | 79 | 24.49 | 3 |
| Italy | 2/20 | 14 | 5 | 15 | 127 | 55.3 | 1 |
| Latvia | 3/7 | 6 | NA | 15 | 104 | 35.59 | 2 |
| Lithuania | 3/10 | 4 | 1 | 17 | 105 | 35.17 | 3 |
| Malaysia | 2/26 | 16 | 5 | 31 | 123 | 17.79 | 3 |
| Malta | 4/1 | -19 | NA | 5 | 99 | 12.72 | 4 |
| Mexico | 3/10 | 13 | 10 | 17 | 38 | 33.13 | 1 |
| Moldova | 3/9 | 2 | 14 | 18 | 36 | 33.58 | 1 |
| Morocco | 3/9 | 7 | 4 | 20 | 78 | 30.97 | 1 |
| Netherlands | 2/28 | 11 | NA | 15 | 126 | 49.74 | 1 |
| New Zealand | 3/13 | 10 | 3 | 16 | 69 | 32.66 | 4 |
| Norway | 2/29 | 12 | NA | 19 | 134 | 27.81 | 3 |
| Poland | 3/5 | 5 | 14 | 18 | 121 | 39.56 | 1 |
| Portugal | 3/3 | 13 | 3 | 24 | 149 | 32.28 | 2 |
| Romania | 3/3 | 3 | 17 | 24 | 85 | 29.23 | 1 |
| S Korea | 2/15 | 16 | NA | 14 | 73 | 44.05 | 4 |
| Serbia | 3/9 | 7 | 12 | 18 | 78 | 37.55 | 1 |
| Slovakia | 3/7 | 5 | NA | 8 | 82 | 61.33 | 1 |
| Slovenia | 3/7 | 5 | NA | 9 | 71 | 43.87 | 2 |
| South Africa | 3/7 | 11 | 8 | 15 | 32 | 43.98 | 1 |
| Spain | 2/24 | 17 | 2 | 26 | 125 | 36.92 | 2 |
| Sweden | 2/25 | 15 | NA | 23 | 183 | 33.49 | 1 |
| Switzerland | 2/26 | 2 | NA | 21 | 96 | 39.02 | 2 |
| Turkey | 3/12 | 4 | NA | 14 | 79 | 65.48 | 1 |
| Ukraine | 3/16 | -4 | NA | 21 | 68 | 29.79 | 1 |
| United Kingdom | 2/25 | 24 | 3 | 36 | 125 | 21.79 | 2 |

**Table Appendix C.2:** COVID-19 case counts; US states

| | day<sub>0</sub> | $t_{\text{policy1}}$ | $\Delta_{\text{lock}}$ | $t_{\text{h}}$ | $t_{\text{end}}$ | $g_{\text{u}}$ | $k$ |
| --- | --- | --- | --- | --- | --- | --- | --- |
| Arkansas | 3/17 | 0 | NA | 5 | 46 | 49.58 | 1 |
| California | 2/27 | 13 | 8 | 36 | 95 | 20.87 | 1 |
| Colorado | 3/8 | 9 | 9 | 18 | 93 | 33.66 | 1 |
| Connecticut | 3/8 | 4 | NA | 19 | 112 | 43.32 | 1 |
| Delaware | 3/14 | 2 | 8 | 18 | 159 | 30.09 | 1 |
| District of Columbia | 3/13 | 0 | 17 | 19 | 108 | 27.31 | 1 |
| Florida | 3/2 | 15 | 17 | 27 | 52 | 32.8 | 1 |
| Georgia | 3/5 | 13 | 16 | 23 | 66 | 34.05 | 1 |
| Idaho | 3/16 | 7 | 2 | 16 | 49 | 31.5 | 3 |
| Illinois | 3/5 | 8 | 8 | 26 | 117 | 33.05 | 1 |
| Indiana | 3/11 | 1 | 13 | 20 | 97 | 36.15 | 1 |
| Iowa | 3/21 | -4 | NA | 15 | 23 | 20.42 | 2 |
| Kansas | 3/13 | 4 | 13 | 23 | 80 | 25.08 | 1 |
| Kentucky | 3/14 | 2 | NA | 19 | 27 | 23.52 | 2 |
| Louisiana | 3/11 | 2 | 10 | 20 | 72 | 33.03 | 2 |
| Maine | 3/12 | 4 | 17 | 8 | 145 | 63.17 | 1 |
| Maryland | 3/7 | 9 | 14 | 29 | 102 | 27.61 | 1 |
| Massachusetts | 3/4 | 9 | NA | 31 | 181 | 28.31 | 2 |
| Michigan | 3/15 | -2 | 11 | 15 | 86 | 38.11 | 2 |
| Minnesota | 3/7 | 10 | 11 | 13 | 23 | 46.06 | 1 |
| Mississippi | 3/14 | 5 | 15 | 11 | 18 | 44.44 | 2 |
| Missouri | 3/11 | 12 | 14 | 16 | 58 | 46.78 | 1 |
| Nebraska | 3/13 | 3 | NA | 53 | 97 | 11.41 | 2 |
| Nevada | 3/7 | 9 | 15 | 19 | 58 | 35.07 | 1 |
| New Hampshire | 3/12 | 4 | 11 | 14 | 155 | 32.9 | 1 |
| New Jersey | 3/5 | 11 | 5 | 26 | 132 | 37.8 | 2 |
| New York | 3/2 | 10 | 10 | 23 | 170 | 45.56 | 2 |
| North Carolina | 3/13 | 1 | 16 | 16 | 33 | 33.24 | 1 |
| North Dakota | 3/16 | 0 | NA | 12 | 69 | 40 | 1 |
| Ohio | 3/10 | 2 | 11 | 16 | 28 | 43 | 1 |
| Oklahoma | 3/12 | 5 | NA | 15 | 72 | 38.9 | 1 |
| Oregon | 3/6 | 6 | 11 | 18 | 75 | 29.72 | 1 |
| Pennsylvania | 3/7 | 10 | 15 | 29 | 98 | 28.87 | 2 |
| Rhode Island | 3/15 | 1 | 12 | 31 | 106 | 18.67 | 2 |
| South Carolina | 3/14 | 2 | 22 | 15 | 34 | 32.3 | 1 |
| South Dakota | 4/2 | -17 | NA | 16 | 98 | 16.64 | 2 |
| Tennessee | 3/12 | 8 | 13 | 15 | 32 | 35.94 | 1 |
| Texas | 3/3 | 16 | 14 | 35 | 77 | 22.31 | 2 |
| Utah | 3/9 | 7 | NA | 16 | 61 | 37.09 | 1 |
| Vermont | 3/20 | -7 | 11 | 11 | 50 | 22.99 | 3 |
| Virginia | 3/7 | 8 | 15 | 15 | 24 | 41.13 | 1 |
| Washington | 2/27 | 13 | 12 | 20 | 192 | 31.64 | 1 |
| Wisconsin | 3/8 | 9 | 8 | 19 | 163 | 37.03 | 1 |

**Table Appendix C.3:** COVID-19 death counts; Nations

| | day<sub>0</sub> | $t_h$ | $t_{end}$ | $g_u$ | $k$ |
| --- | --- | --- | --- | --- | --- |
| Argentina | 3/23 | 10 | 69 | 27.67 | 1 |
| Austria | 4/2 | 14 | 105 | 7.79 | 8 |
| Belgium | 3/13 | 24 | 164 | 28.94 | 3 |
| Bulgaria | 4/18 | 116 | 150 | 2.72 | 1 |
| Canada | 3/21 | 33 | 165 | 15.82 | 3 |
| Colombia | 3/31 | 14 | 22 | 17.81 | 3 |
| Croatia | 4/1 | 13 | 58 | 15.35 | 2 |
| Czechia | 3/25 | 13 | 99 | 23.49 | 3 |
| Denmark | 3/15 | 19 | 149 | 24.13 | 3 |
| Dominican Republic | 3/21 | 6 | 292 | 57.31 | 1 |
| Ecuador | 3/18 | 22 | 276 | 24.04 | 1 |
| Egypt | 3/17 | 48 | 204 | 10.64 | 1 |
| Estonia | 4/11 | 6 | 106 | 8.48 | -1 |
| Finland | 3/24 | 19 | 106 | 21.9 | 2 |
| France | 3/5 | 31 | 137 | 23.2 | 4 |
| Germany | 3/12 | 22 | 123 | 28.48 | 2 |
| Hungary | 4/4 | 15 | 105 | 14.29 | 3 |
| Ireland | 3/21 | 18 | 126 | 26.68 | 2 |
| Israel | 3/23 | 11 | 83 | 35.6 | 2 |
| Italy | 2/25 | 28 | 172 | 25.01 | 3 |
| Malaysia | 3/19 | 5 | 64 | 52.59 | 2 |
| Mexico | 3/21 | 21 | 23 | 27.87 | 1 |
| Moldova | 3/31 | 12 | 139 | 22.79 | 1 |
| Morocco | 3/21 | 13 | 83 | 26.18 | 3 |
| Netherlands | 3/13 | 20 | 135 | 26.07 | 3 |
| New Zealand | 4/9 | 5 | 55 | 41.81 | 4 |
| Panama | 3/21 | 6 | 67 | 50.88 | 1 |
| Peru | 3/26 | 39 | 280 | 14.33 | 1 |
| Philippines | 3/11 | 21 | 295 | 23.57 | 1 |
| Portugal | 3/17 | 14 | 136 | 37.99 | 2 |
| Romania | 4/25 | 19 | 35 | 3.6 | 8 |
| Serbia | 4/8 | 12 | 43 | 9.05 | -1 |
| Singapore | 4/1 | 22 | 79 | 7.49 | -1 |
| Slovenia | 3/21 | 13 | 72 | 25.6 | 2 |
| South Africa | 5/14 | 81 | 161 | 5.12 | -1 |
| Spain | 3/3 | 22 | 82 | 38.38 | 2 |
| Sweden | 3/13 | 22 | 164 | 26.29 | 2 |
| Switzerland | 3/16 | 20 | 212 | 19.3 | 6 |
| Turkey | 3/18 | 18 | 140 | 36.51 | 2 |
| Ukraine | 4/17 | 29 | 45 | 6.53 | -1 |
| United Kingdom | 3/11 | 23 | 156 | 29.97 | 2 |

**Table Appendix C.4:**
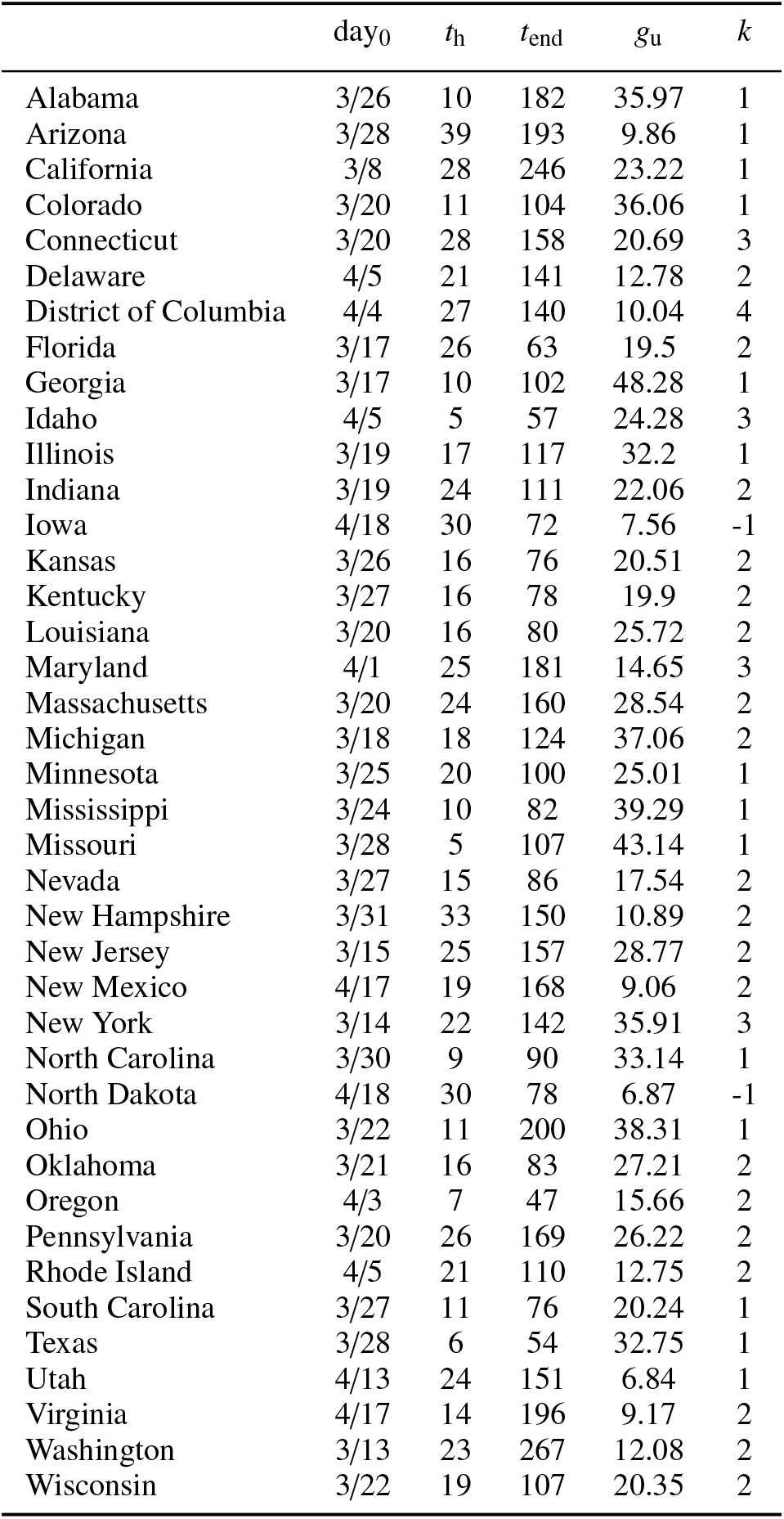
COVID-19 death counts; US states

## Footnotes

1 Negative *a*_*k*_ describe accelerated growth (*dg*/*dt* > 0), which we do not consider here.

2 http://www.healthdata.org/

3 Pandemic data for nations were taken from the Johns Hopkins University COVID-19 Data Repository https://github.com/CSSEGISandData/COVID-19, for US states from the New York Times site https://github.com/nytimes/covid-19-data.

4 The t-statistic associated with this estimate is 1.30, hence the two-sided p-value of 0.20. Thus, the 80% confidence interval has a lower bound of 0. This implies, assuming a symmetric distribution of the coefficient estimate, that approximately 10% of the distribution density falls below zero.

5 There is less information on the ordering of policy implementation in specific regions in China. They do note, though, that policies were deployed there in a “centralized manner” between January 23–29, 2020, and suggest that the first policy deployed locally was a “level 1 emergency declaration,” which does not include lockdown.

6 For a detailed description of the MK-test see Kocsis et al. (2017).

7 The *F*-test is closely related to the *odds ratio* test in Bayesian statistics. The two become the same if and only if one assumes scale-invariant Jeffreys’ prior for RSS (Ivezić et al., 2014).

